# SARS-CoV-2 surveillance in untreated wastewater: first detection in a low-resource community in Buenos Aires, Argentina

**DOI:** 10.1101/2020.10.21.20215434

**Authors:** Néstor Gabriel Iglesias, Leopoldo Germán Gebhard, Juan Manuel Carballeda, Ignacio Aiello, Emiliano Recalde, Gabriel Terny, Silvina Ambrosolio, Gabriela L’Arco, Jonatan Konfino, Juan Ignacio Brardinelli

## Abstract

Recently, the World Health Organization (WHO) in a brief report about the status of environmental surveillance for SARS-CoV-2 indicated that *approaches are needed that can be applied in lower-resource settings, where a greater proportion of the population is not connected to sewers and instead uses pit toilets or septic tanks. Possibilities include testing surface water contaminated by sewage*. In this study we measured SARS-CoV-2 RNA from a surface water source in a low-income settlement. We observe for this community that measurements of SARS-CoV-2 concentrations in surface water contaminated by sewage can be considered as an estimation of changes in COVID-19 prevalence on a population level.

## Introduction

Wastewater-based epidemiology (WBE) provides comprehensive health information at the community level [1]. The concept is mainly based on the detection and analysis of chemical and biological compounds in sewage. WBE is an approach used to monitor the presence of pathogens which may pose a public health concern [2]. During the current COVID-19 pandemic, sewage surveillance by analyzing for SARS-CoV-2 RNA traces in wastewater has been reported in many locations around the world. All these reports have been conducted in populations which have sewer networks and wastewater treatment facilities [3-6]. However, no study has been reported with this approach in low-resource settlements lacking these facilities.

On January 10^th^ 2020 the World Health Organization (WHO) published a comprehensive package of guidance documents for countries, covering topics related to the management of an outbreak of a new disease, including recommendations about surveillance [7]. On March 11^th^, the WHO declared COVID-19 as a pandemic [8], and on March 18^th^, the first case was confirmed in the Municipality of Quilmes, Province of Buenos Aires, Argentina and the prevention and control strategy of COVID-19 began to be implemented [9]. On May 25^th^ a COVID-19 outbreak in the Villa Azul neighborhood of Quilmes was confirmed and an outbreak mitigation strategy was implemented in addition to a comprehensive surveillance strategy that also was extended to Villa Itatí community given the geographical and social proximity [10].

In this scenario, a surveillance strategy was developed in Villa Itatí including an active search of eventual new cases (suspected cases) house by house, the registry of new COVID-19 cases [10] and wastewater surveillance. Every suspected case under the current epidemiological guidelines was tested with a nasal and oral swab for real time PCR analysis. Once the case was confirmed, isolation measures were recommended for the recovery and avoidance as much as possible more contagiousness and more cases [10].

The aim of this study was to analyze the presence of SARS-CoV-2 RNA in surface waters contaminated with wastewater from a low-income settlement and to analyze its relationship with the prevalence of COVID-19 disease at the population level as part of the surveillance strategy.

## Material and methods

Raw surface water samples were collected between June 5^th^ and September 7^th^ at Villa Itatí neighborhood, Municipality of Quilmes, Buenos Aires, Argentina. It is estimated that Villa Itatí has a population of 16,478 people, in an area of 55 hectares delimited by Montevideo, Levalle, Ayacucho and Southeast access streets [11]. It is a neighborhood that has an average of 1.03 households per dwelling with on average 3.55 people [12]. Of the 4,261 homes in Villa Itatí, 3,966 (93.1%) have potable water from the network and 1,044 (24.5%) are connected to the public sewer network [12].

This community is characterized by being located around an endorheic urban basin formed by an anthropogenic digging flooded by the groundwater table and the discharges of pluvial and domestic waters [13]. Most households serve their sewage into the waterlogged digging (lagoon) through open drains or indirectly by underground infiltration from pit toilets and cesspools. In order to avoid the overflowing of the lagoon, its content is daily discharged to the urban rainwater network through a pumping system [13]. Water samples were taken from the unique pumping station of the neighborhood.

Composite samples representing 6-hour period (from 8 a.m. to 2 p.m.) were collected in sterilized 500 ml glass bottles and kept at 4 °C until analysis, the RNA purification and viral detection were performed the same day as the sample was collected. Before processing, samples were subjected to a 90 minutes treatment at 60 °C to ensure biological safety. Viral concentration was carried out from 250 ml of sample adding 20 grams of PEG_8000_ and 4.5 grams of NaCl and then centrifuging at 12.000 g during 1 hour at 4°C. The pellet was suspended in 1 ml of Trizol (Invitrogen) and RNA was purified according to the supplier´s protocol. The extracted RNA was further purified using QiAamp Viral RNA Mini Kit from QIAGEN and eluted in 50 ul of ribonuclease-free water. SARS-CoV-2 RNA was quantified by RT-qPCR using the CDC N1 and N2 probe/primers sets obtained from Integrated DNA Technologies Inc. An RT-qPCR assay targeting pepper mild mottle virus (PMMoV) was performed as viral indicator of human fecal contents [4, 14]. Samples were analyzed using GoTaq Probe 1-Step RT-qPCR System (Promega) in 20 ul reaction mix in a QuantStudio 3 Real-Time PCR System (Applied Biosystems). In our conditions, we obtained a standard curve for N1 primer set with an R^2^ of 0.99 with efficiency of 85% (slope=-3.747; y intercept = 46.076). The N2 primer set generated a standard curve with an R^2^ of 0.98 with an efficiency of 92.3% (slope=-3.508; y intercept=42.014). All determinations were made at least in duplicate.

## Results

We collected raw water samples eleven times from June to September at Villa Itatí pumping station. Sampling was performed weekly with some exceptions mainly due to impediments caused by heavy rains.

We optimized an RNA isolation procedure consisting in a PEG_8000_ based precipitation step in order to achieve viral concentration from the raw water samples. Then, a Trizol (chaotropic agents/organic solvents) extraction followed by a silica-gel based RNA purification step were carried out in order to recover the viral RNA free from interfering substances.

Real time RT-qPCR was performed using the same CDC N1 and N2 probe/primers sets which are utilized for COVID-19 diagnostic [15]. We detected SARS-CoV-2 RNA in all the samples, and also, we were able to quantify the relative SARS-CoV-2 RNA concentration in these samples using both N1 and N2 sets. The RT-qPCR cycle threshold values (Ct) ranged from 32 to 40 with means of 35,4 and 34,4 for N1 and N2, respectively. The comparison of the relative RNA concentrations obtained by the N1 and N2 sets is shown in Figure 3. The correlation exhibited a R^2^ of 0.84 (Figure 3). In all samples we also determined the relative amount of PMMoV RNA as a viral indicator of human feces content, obtaining Ct values ranged from 30 to 33 with an means of 31,9.

**Figure 1.**
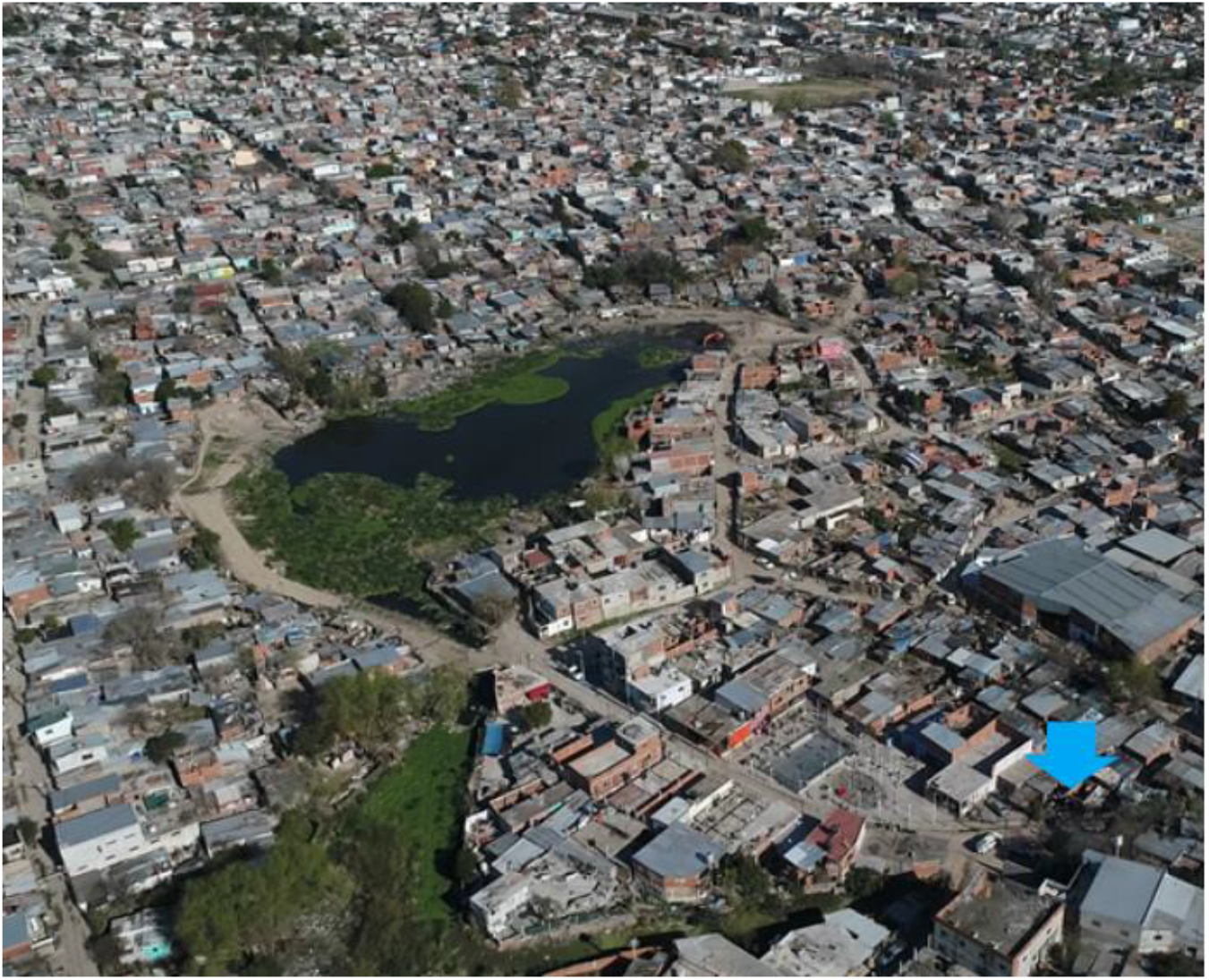
Aerial view of Villa Itatí neighborhood, Quilmes, Buenos Aires, Argentina with the lagoon and the pumping station indicated (arrow). Source: Ariel Romaniuk, Municipality of Quilmes.

**Figure 2.**
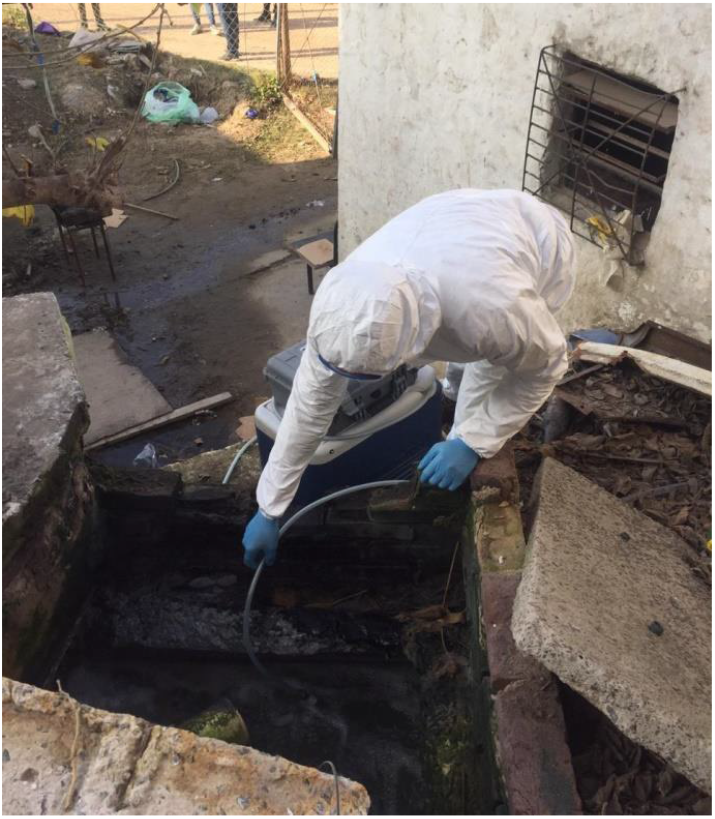
An OPDS worker taking a water sample at Villa Itatí pumping station. Source: OPDS.

**Figure 3.**
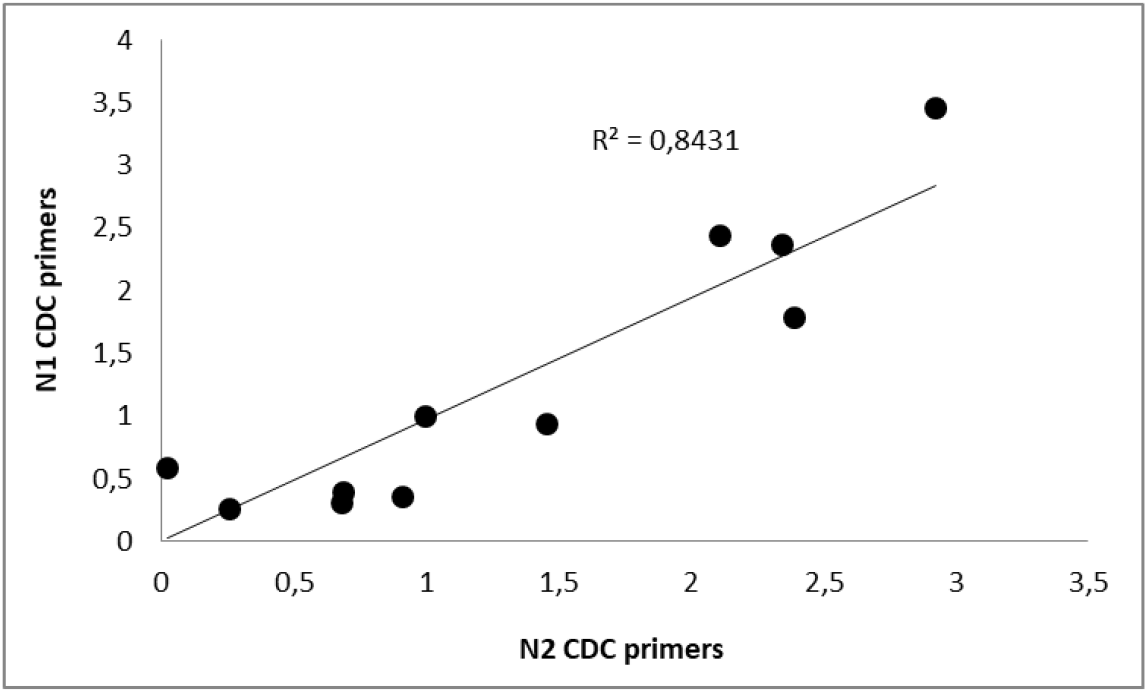
Comparison of SARS-CoV-2 relative RNA concentrations measured in surface water samples using CDC N1 and N2 probe/primers sets.

In order to determine the number of active COVID-19 cases in the Villa Itatí population for each date, we assumed an active case beginning from the date of positive diagnosis for COVID-19 until the next 15 days, since it is the median period of persistent viral shedding of SARS-CoV-2 in feces and urine [16, 17]. We obtained a curve of active cases for this population based on data provided by the Secretary of Health from the Municipality of Quilmes. We observed an increase in the number of cases from May 28^th^ until reaching a plateau on June 13^th^ that remained stable until July 1^st^ and then began to decline until July 16^th^ when a new increase began until reaching the maximum number of cases on August 11^th^ when the cases began to decline until the end of the date of this work on September 24^th^.

The presence of SARS-CoV-2 genetic material was detected at the time when the reported cases in the neighborhood were 122 over 16.478 inhabitants in June 5 (Figure 4), indicating that a sensitivity of less than 1 reported case per 135 inhabitants could be achieved in this particular setting.

**Figure 4.**
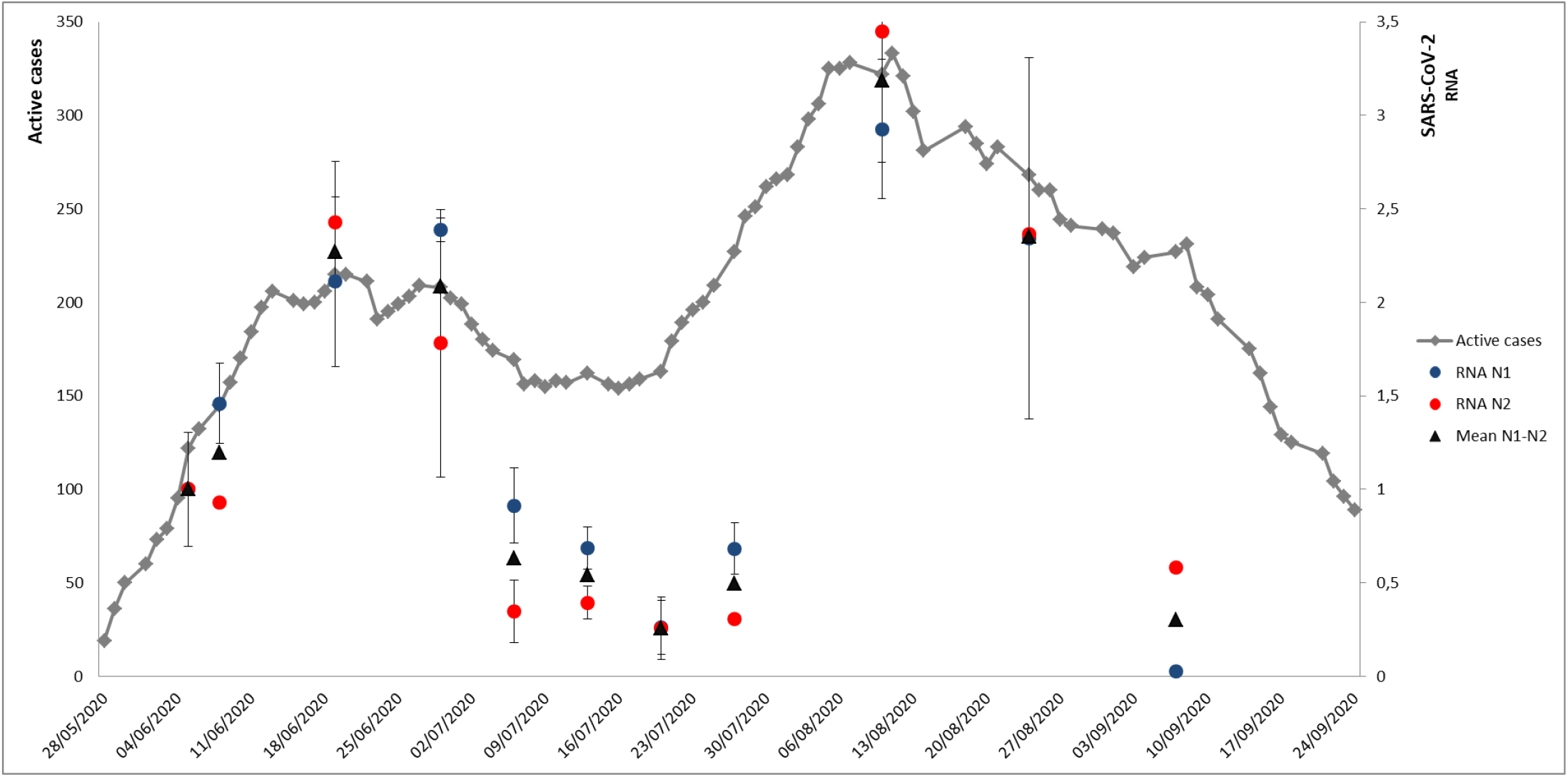
Determinations of relative SARS-CoV-2 RNA concentration and active COVID-19 cases per day. Diamonds (grey) indicates the number of active cases. The relative RNA of SARS-CoV-2 for each set of primers is indicated by circles (N1 in blue and N2 in red). The mean of relative RNA concentration is represented with triangles. All measurements were made in duplicate, error bars indicate the standard deviation.

The results are summarized in Figure 4 as the mean of the relative RNA concentration for N1 and N2 probe/primers sets. We detected that the change in the SARS-CoV-2 RNA concentration in the water samples obtained for both N1 and N2 matched the trend observed for the number of COVID-19 active cases along time. In other words, the relative SARS-CoV-2 RNA concentration determined in environmental samples traced the shape of infections during the analyzed period.

We observed a good correlation between the concentration of viral RNA found in the wastewater samples and the number of active cases reported by the health system. We were able to corroborate that as the positive cases increased in the settlement, a higher concentration of viral RNA in the sewage was observed, while when the positive cases decreased after the outbreak mitigation process was implemented and social distancing measures addressed, lower concentrations of viral RNA were detected in the wastewater samples.

## Discussion

Most studies published to date on the use of environmental surveillance for SARS-CoV-2 have been from high income settings [18]. This is the first report of SARS-CoV-2 RNA detection from a surface water source of a marginalized population with a precarious wastewater drainage. Additionally, to our knowledge, it is the first experience of utilizing SARS-CoV-2 RNA measurement in wastewater as environmental surveillance strategy for the COVID-19 pandemic in Argentina.

Nevertheless, we need to acknowledge some limitation. The strategy has some restrictions due to the aforementioned characteristics regarding the neighborhood infrastructure and the sampling; the hydraulic retention time has to be determined and sampling more frequently may reduce the variance and noise in the data. There are also uncertainties due to limited knowledge about SARS-CoV-2 gastrointestinal infection, the concentration of viral RNA in stool over the course of the illness, the variability in viral dynamics in individuals and fecal shedding [16, 17, 19, 20, 21]. Because of this, the number of cases cannot be estimated with confidence directly from the quantification of viral RNA in wastewater [19]. However, we were able to detect the presence of SARS-CoV-2 genetic material and follow the dynamics of the infection in the community. We believe that the methodology could be improved by increasing the sampling frequency and time of sampling. Although this methodology has its limitations, lengthwise trends of SARS-CoV-2 RNA concentration in fecal contaminated surface water may still be useful in supplementing conventional surveillance methods to interpret the trends in community transmission.

A more comprehensive COVID-19 surveillance system could be developed if wastewater SARS-CoV-2 frequent measurement is included, taking into account the disposal, fate, and transport of wastewater in each community. Sewage testing has been successfully used as a method for early detection of other pathogens, such as Poliovirus [22]. Since SARS-CoV-2 can be shed in the feces of individuals with symptomatic or asymptomatic infection wastewater surveillance can capture data on both types of infection [16, 20, 21]; it can be a leading indicator of changes in COVID-19 burden in a community; and could serve as a COVID-19 indicator that is independent of healthcare-seeking behaviors and access to clinical testing [19].

Overall, our results demonstrate for this community that measurements of SARS-CoV-2 RNA levels in surface water contaminated by sewage can be considered as a reliable estimation of changes in COVID-19 prevalence on a population level. This method may be applied to detect outbreaks or an increase in the number of infected cases and is a useful complement to classic epidemiological surveillance to assess whether the measures applied are effectively working.

## Data Availability

All relevant data is given in the manuscript

## Funding

Programa de articulación y fortalecimiento federal de las capacidades en Ciencia y Tecnología COVID-19, from the Ministry of Science, Technology and Innovation of Argentina.

**The authors declare no conflict of interests.**

## Acknowledgements

We wish to thank the OPDS laboratory staff for the sampling task and Carolina Vera from the Ministry of Science, Technology and Innovation of Argentina for her support to the project.

## Notes

### Competing Interest Statement

The authors have declared no competing interest.

### Funding Statement

Funding: Programa de articulacion y fortalecimiento federal de las capacidades en Ciencia y Tecnologia COVID-19, from the Ministry of Science, Technology and Innovation of Argentina.
Acknowledgements: We wish to thank the OPDS laboratory staff for the sampling task and Carolina Vera from the Ministry of Science, Technology and Innovation of Argentina for her support to the project. 

### Author Declarations

No oversight was required

